# Rural and urban disparities in malnutrition and quality of life among older persons in Lagos Nigeria: A comparative analytical cross-sectional study

**DOI:** 10.1101/2025.08.01.25332605

**Authors:** Babatunde Abdulmajeed Akodu, Monsuru Owolabi Badmus, Gbenga Olorunfemi, Oluchi Joan Kanma-Okafor, Olufunmilayo Abeni Olokodana-Adesalu, Moninuola Seliat Ojikutu, Taiwo Hussein Agunbiade

## Abstract

**Background:** Malnutrition has been identified as an adverse condition that affects the health of older persons. Despite being recognized as one of the leading causes of disease in older persons, very little attention has been given to malnutrition among older persons. Different studies carried out on nutritional status and quality-of-life among older persons in rural and urban areas have shown disparities in their results. The aim of this study was to compare the nutritional status and quality-of-life of the elderly living in rural and urban areas of Lagos State.

**Method:** This was an analytical cross-sectional study involving 331 participants for nutritional assessment using a Mini-nutritional assessment short form while quality-of-life was assessed using the WHOQOL-AGE. Data was analysed using STATA 13.0 software. Association between variables was determined and multiple logistic regression model was used to determine the predictor of malnutrition and quality-of-life.

**Results:** 67.7% of the participants in rural area were at risk of malnutrition while 49.4% were at risk of malnutrition in urban area. The predictors of malnutrition in rural area were age (OR: 1.1, 95%CI: 1.0 – 1.2, P < 0.01), and quality-of-life (OR: 0.2, 95%CI: 0.07 – 0.58, P < 0.003) while age (OR: 1.1, 95%CI: 1.01 – 1.16, P < 0.01), quality-of-life (OR: 0.29, 95%CI: 0.1 – 0.9, P < 0.03), sex (OR: 3.3, 95%CI: 1.04 – 10.5, P < 0.01), education level (OR: 2.05, 95%CI: 1.02 – 4.16, P < 0.04), and income (OR:0.21, 95%CI: 0.04 – 0.93, P < 0.04) were predictors of malnutrition in urban area while nutritional, sex and marital status were predictors of quality-of-life.

**Conclusion and Recommendation:** Significant proportion of older persons in both rural and urban area were at risk of malnutrition while sex, quality-of-life, education level, and income were predictors of malnutrition and awareness campaign on prevention strategies for malnutrition on the elderly is vital.

## Introduction

An urban area has been described as the region surrounding a city where most inhabitants of those areas have non-agricultural jobs. [1]When compared with urban areas, rural area is described as the opposite of urban area where their dwellers are generally less wealth, with less access to medical services, low levels of education and less organized communities in terms of housing planning in which these factors can affect their Quality of Life (QoL) and their health status [2]

There are inconsistencies on the predictive parameters for nutritional status and quality of life among elderly in rural and urban areas. It was revealed by several studies that the elderly individuals living in rural areas have lower QoL and exhibits worse health conditions than elderly individuals living in urban areas[3] Although a study also reported lower QoL among urban dwellers.[4] The lower QoL reported by some studies among rural dwellers was associated with their poor health behaviour which also include their eating behaviour.[3] Unhealthy eating behaviour have been reported by several studies as a factor that affects the elderly nutritional status and make them be at risk of malnutrition.[5]. In developing countries including Nigeria most elderly moved into old age facing challenges of continuing poverty, inadequate access to good health, deprivation and poor diet in term of quality and quantity.[6] Malnutrition in elderly can be described in term of under nutrition and overweight.[6,7]. In Nigeria, a study among the low income area of southwest showed that 18% of aged that participated in that study were malnourished while another study in the rural communities in Osun State revealed that about 35% of the respondents were in risk of malnutrition.[5] A study among the older person presenting in a primary care clinic in urban area of Nigeria also showed that 61.7% of the elderly have nutritional problem.[8]. In Nigeria, there is a limited study involving alternative measures of nutritional status and QoL among rural and urban dweller elderly and how their place of residence impact their nutritional status. This makes it important for researchers to study more about the Quality of Life and nutritional status of this vulnerable group in respect of where they resided. Also, Dewi et al reported 67% of elderly have high risk of malnutrition in the study among the elderly living in rural areas of India.[9] In Africa, 58.5% of elderly were reported to be at risk of malnutrition among urban dweller in Niamey.[10]. A study among elderly living in rural and urban area in Ethiopia using MNA reported that 4.9% were malnourished in urban area while 17.1% of the elderly in rural area were malnourished.[11]. In another study among the rural dwellers elderly in Osun State, it was reported that 35% of the elderly were at risk of malnutrition[5] while Adebusoye et al reported that 11.8% of the elderly were at risk of malnutrition in the study conducted among the elderly in urban areas in southwestern Nigeria.[8]. It was revealed that living in rural areas have 2.24 odds of been at risk of malnutrition than living in urban area.[11]. A study of QoL among the elderly in rural populace in Nigeria reported that over average of the respondents have good quality of life and age, gender, marital status, occupation and income were statistically significant with the QoL.[12] Oladipupo et al also reported that the urban dwellers elderly (31.5%) have higher good QoL than that of rural dwellers (20.5%) in a study conducted among the elderly in Ilesa. It also reported that those who have good family support have higher quality of life than those who does not.[4]. This study aimed to compare the nutritional status and quality of life of the elderly living in rural and urban areas of Lagos State

## Materials and Method

Study area: Lagos State is located on the South–Western part of Nigeria and covered an area of 358,862 hectares or 3,577 sq. km. which represents 0.4% of Nigeria’s territorial land mass of 923,773 sq. km. Lagos State is the smallest state in Nigeria yet, it has the highest urban population, which is 27.4 % of the national estimate.[13] According to the 2006 National Census, Lagos State has a population of 9,013,534 in relation to the National count of 140,003,542 with the elderly population of 331,071.[14] However, based on the UN-Habitat and international development agencies’ estimates, Lagos State is said to have about 24.6 million inhabitants in 2015.[13]Lagos State is divided into 20 Local Governments and 37 Local Council Development Areas respectively, in accordance with Nigeria’s federal structure and the need to bring governance, development and participatory democracy to the grassroots. 16 out of the Local Government were in urban area while the remaining 4 were in rural area rural.[15,16] The local government areas in urban area include; Surulere, Ajeromi/Ifelodun, Agege, Ojo, Apapa, Lagos Mainland, Mushin, Alimosho, Ifako-Ijaye, Ikeja, Lagos Island, Kosofe, Eti-Osa,Amuwo Odofin, Oshodi/Isolo, Shomolu while the local government areas in the rural area includes; Ikorodu, Epe, Ibeju-Lekki and Badagry.[15,16] The elderly populations were widely distributed across all the Local Government Areas in the State.

### Study design

This study is a comparative analytical cross-sectional study that assessed the quality of life and nutritional status of the elderly in rural and urban of Lagos State and This study involved the elderly living in Badagry and Mushin Local Government Area of Lagos State.

### Eligibility Criteria

Inclusion criteria. Elderly[17] who gave consent and over 60 years of age were included. Exclusion criteria. Elderly that have any form of disability were excluded from the study.[18] Also, elderly who decline to participate in the study were excluded.

Sample size determination

The minimum sample size was estimated using the following formula[19]

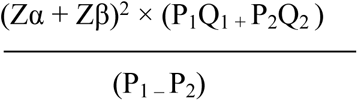

Where Zα = Confidence level 95% or 0.05 or 1.96, Zβ = Power of 0.10 = 1.28, P_1_ = prevalence of malnutrition among elderly living in rural area[5] 0.35, P_2_ = prevalence of malnutrition among elderly living in urban area[8] 0.617, Q_1_ = 1 - P_1_ and Q_2_ = 1 – P_2._ An estimated sample size of 154 was gotten and upgraded by adding 10% attrition to obtain a final estimate of 170 for each group

### Sampling technique and data collection tools

The target population for the study were male and female elderly that resided in the study area and at least 60 years of age. A multistage sampling technique was used in selection of participants for the study from 15^th^ June to 18^th^ July 2021.

#### First stage

Out of 20 local Government Area in Lagos State (16 Urban and 4 Rural)[15,16], one Rural and one Urban were randomly selected using simple balloting without replacement.

#### Second stage

Based on available information [15,16], rural area ward usually contains 12 streets with 40 houses and each houses contains 3 or more households while urban area wards usually contains 17 streets with 60 houses each and each houses usually contains 4 or more households. Based on this, one ward each were selected using simple random technique in the selected Rural and Urban Local overnment Area.

#### Third stage

6 streets each were randomly selected from the selected ward in the rural and urban area.

#### Third stage

In this last stage, participants that meet the inclusion criteria were recruited from each household in the houses that were located in the 6 selected streets consecutively until the sample size is reached. Where a household declined, the next household was recruited.

### Method of data collection

The data for this study was collected using a well-structured questionnaire that was developed from past studies. The questionnaire was divided into four sections which are;

#### Section A

Socio-demographic data of the respondents. These are the socio-demographic of the respondents that were adopted from the similar studies [8]

#### Section B

Nutritional status of respondents using Mini nutritional assessment short form (MNA-SF). This tool has been used in a previous study in Nigeria [5,8] to check the nutritional status of elderly. In 2009 a Revised MNA-SF was developed & validated as a stand-alone screening tool; it includes option for substituting calf circumference (CC) for BMI, and takes less than 5 minutes.[20] MNA-SF have been validated in many studies. [21,22] The sensitivity and specificity of the MNA-SF with a sensitivity of 87% and a specificity of 85% against the MNA, and a sensitivity of 81% and a specificity of 63 % against a wide range of criteria in studies[23]. MNA-SF is appropriate screening and assessment tools for use in community-dwelling elderly, and all other geriatric settings [24]

#### Section C

Determined respondents’ quality of life using WHOQOL-AGE. This tool has been validated by different studies [25,26] The WHOQOL-AGE shows promising psychometric properties in nationally representative with Cronbach’s alpha was 0.88 for factor 1, and 0.84 for factor 2.[25] The convergent validity of the WHOQOL-AGE was estimated at 0.75 [95% CI = (0.73, 0.77)].[25] The 13-item WHOQOL-AGE was designed according to the characteristics of aging populations. Four research assistants with school certificate as the minimum education who were proficient in English, Yoruba and Pidgin languages was recruited and trained to help with the collection of data from the respondents. All activities of the research assistants were monitored and supervised properly. Also, research assistant translated questionnaire to language understood by the respondents for those that did not understand the English language used to design the questionnaire.

Anthropometric measurement that was collected includes weight and height. The weight of the subjects was measured while standing with both arms by the side and with only light clothing on. The pointer of the weighing scale (Hanson model) was adjusted to zero before each weighing and was recorded to the nearest 0.1kg. In measuring the height of the respondents, wall mount and regular measuring tape was used. The wall mount tape was affixed on the wall; the participant was asked to stand barefoot with heels and toes together. The back of the head, shoulders and buttocks touched the wall. A metallic plate was lowered from the height compressing the hair and finding the top of the head and the reading was measured closest to 0.1cm and height was properly recorded. The body mass index of the elderly was calculated as weight of each individual in kg divided by the square of the height in metres[27].

### Data analysis

Data were entered using Microsoft Excel and was analysed using STATA 13.0 software. Descriptive statistic was computed for all the variables. The independent variables included socio-demographic characteristics (age sex, marrital status,family support, education, income; anthropometric parameters (weight, height and BMI). Dependent variables included Quality of life, place of resisdence (rural/urban), the predictors of nutritional status and Quality of Life included age, family support, sex, marrital status, education. Association between variables was determined using Pearsons Chi-square for categorical variables while Student T-test was used for the numeric variables. Multiple logistic regression model was used to determine the predictor of malnutrition and quality of life. The model was build with variable that have less than P-Value of 0.2 in a univariable regression and some variable were considered APIORI. Level of significance was defined at 95% confidence interval (p<0.05)

### Scoring and grading

The original version of Mini Nutritional Assessment short form (MNA-SF) which was used in this study included six questions. The total score of MNA-SF is graded between Normal nutritional status (score 12-14), at risk of malnutrition (score 8-11) and malnourished ones (score 0-7).

The quality-of-life was measured using WHOQOL-AGE which was graded using likert scale. Summated scores were used to arrive at a quality-of-life score, and the possible ranges of scores are 13-65. The score was categorized as good quality of life (≥40) and poor quality of life (≤39)

### Ethical consideration

Ethical clearance for the study was gotten from the Human Research Ethics Committee (HREC) of the Lagos University Teaching Hospital, Idi-Araba with number ADM/DSCST/HREC/APP/4312. Written informed consent and voluntariness to participation was obtained from all the participants before the interview started. Prior to this, the interviewer explained the purpose of the study, risks, and benefits to participation, the right to withdraw at any time without penalty, and assurance of confidentiality. Privacy was ensured during interviews. No incentives were given for participation.

## Results

A total of 331 (97.4%) from 340 participants recruited for both arms completed the study. A total of 167 of the 170 participants from rural (98.2%) completed the questionnaire while 164 of the 170 participants from urban (96.5%) completed the questionnaire.

Table 1 below depicts the mean age was 65.4±5.5 in rural while it was slightly higher in urban area at 66.1±5.1 and is not statistically significant. The majority of the participants in rural area were female which is slightly above average while most of the participants in urban area were female at 76.8% and is statistically significant with p-value of 0.001. The number of participants that were married is higher in rural than urban with a p-value of 0.005 respectively. Higher percentages of the participants were Yoruba representing 52.1% and 88.4% in rural and urban area respectively with a p-value of 0.001. The percentage of participants that were self employed in both areas were closed which is statistically significant with a p-value of 0.001

**Table 1:** Association between socio demography of the respondents and place of residence.

| Variable | Rural 167<br>Frequency(%) | Urban 164<br>Frequency (%) | P – Value |
| --- | --- | --- | --- |
| Age in years(N)Mean±SD | 65.4±5.5 | 66.1±5.1 | 0.25<sup>b</sup> |
| 60 – 67 | 118(70.7) | 102(62.2) | 0.10 |
| 68 – above | 49(29.3) | 62(37.8) |  |
| Sex |  |  | 0.001* |
| Male | 95 (56.9) | 38(23.2) |  |
| Female | 72 (43.1) | 126(76.8) |  |
| Marital Status |  |  | 0.005* |
| Married | 106(63.5) | 81(49.4) |  |
| Separated | 13(7.8) | 14(8.5) |  |
| Divorced | 26(15.6) | 20(12.2) |  |
| Widowed | 22(13.2) | 50(29.9) |  |
| Ethnicity |  |  | 0.001* |
| Igbo | 16(9.6) | 17(10.4) |  |
| Yoruba | 87(52.1) | 145(88.4) |  |
| Other | 64(38.3) | 2(1.2) |  |
| Religion |  |  | 0.001* |
| Christianity | 81(48.5) | 86(52.4) |  |
| Islam | 52(31.1) | 75(45.7) |  |
| Traditional | 34(20.4) | 3(1.8) |  |
| Level of Education |  |  | 0.001* |
| None | 39(23.4) | 67(40.9) |  |
| Primary | 49(29.3) | 86(52.4) |  |
| Post primary school | 79(47.3) | 11(6.7) |  |
| Occupation |  |  | 0.001* |
| Unemployed | 36(21.6) | 10(6.10) |  |
| Self employed | 108(64.7) | 100(60.9) |  |
| Employed | 23(13.8) | 54(32.9) |  |

Table 2 shows the mean weight of the participants in the rural and the urban area was 63.9±7.8kg and 60.2±12.4kg respectively. Also, the mean weight of the participants in rural and urban area was 1.64±0.1m and 1.61±0.1m respectively. There is a significant association between place of residence and anthropometric measurement (weight and height). Participant mean weight in rural area is higher than urban area at p = 0.001 and they also have higher mean height at p = 0.001.

**Table 2:**
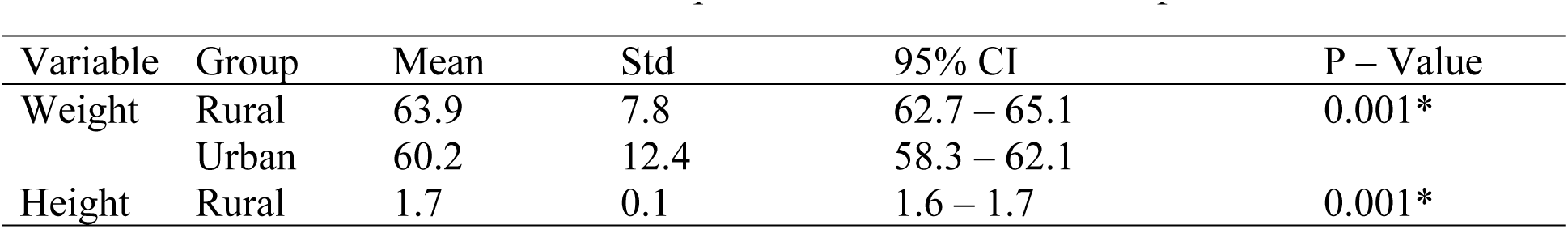
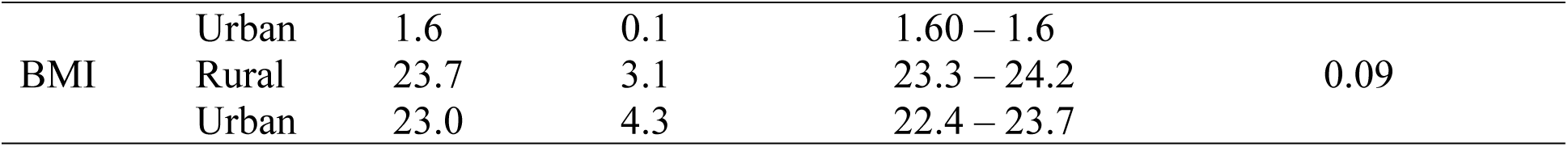
Association between anthropometric measurements and place of residence.

Table 3 revealed that the majority of the participants (67.66) were at risk of malnutrition in the rural area while close to average were at risk of malnutrition in urban area. This is statistically significant with P – value 0.001. Participants at risk of malnutrition were higher in the rural area when compare with the urban area.

**Table 3:** Association between nutritional status of respondents and place of residence.

| Variable | Rural 167<br>Frequency (%) | Urban 164<br>Frequency (%) | P – value |
| --- | --- | --- | --- |
| Malnourished | 23(13.8) | 8(4.9) | 0.001* |
| At risk of Malnutrition | 113(67.7) | 81(49.4) |  |
| Normal Nutrition status | 31(18.6) | 75(45.7) |  |

Pearsons's chi - square

In table 4, there was a statistically significant association between the nutritional score of the participants and their place of residence (P = 0.0008). The mean score of nutritional status is higher in urban area (CI: 10.71 – 11.78)

**Table 4:** Association between nutritional score and place of residence.

| Group | Mean | Std | 95% CI | P – value |
| --- | --- | --- | --- | --- |
| Rural | 10.11 | 2.50 | 9.73 – 10.50 | 0.0008* |
| Urban | 11.24 | 3.45 | 10.71 – 11.78 |  |

Table 5 showed that there was a statistically significant association between the nutritional status and some socio demographic (Age, marital status, level of education, income and occupation) of the respondents in rural area of the participants with P-Value = 0.008,0.007, 0.001,0.01 and 0.001 respectively. Participants who aged between 68 years and above were more at risk of malnutrition and those who were divorced have higher percentage of been malnourished. Majority of those who are malnourished are those who earn less than 30,000 monthly

**Table 5:**
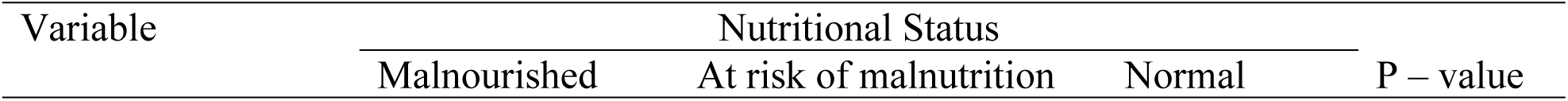
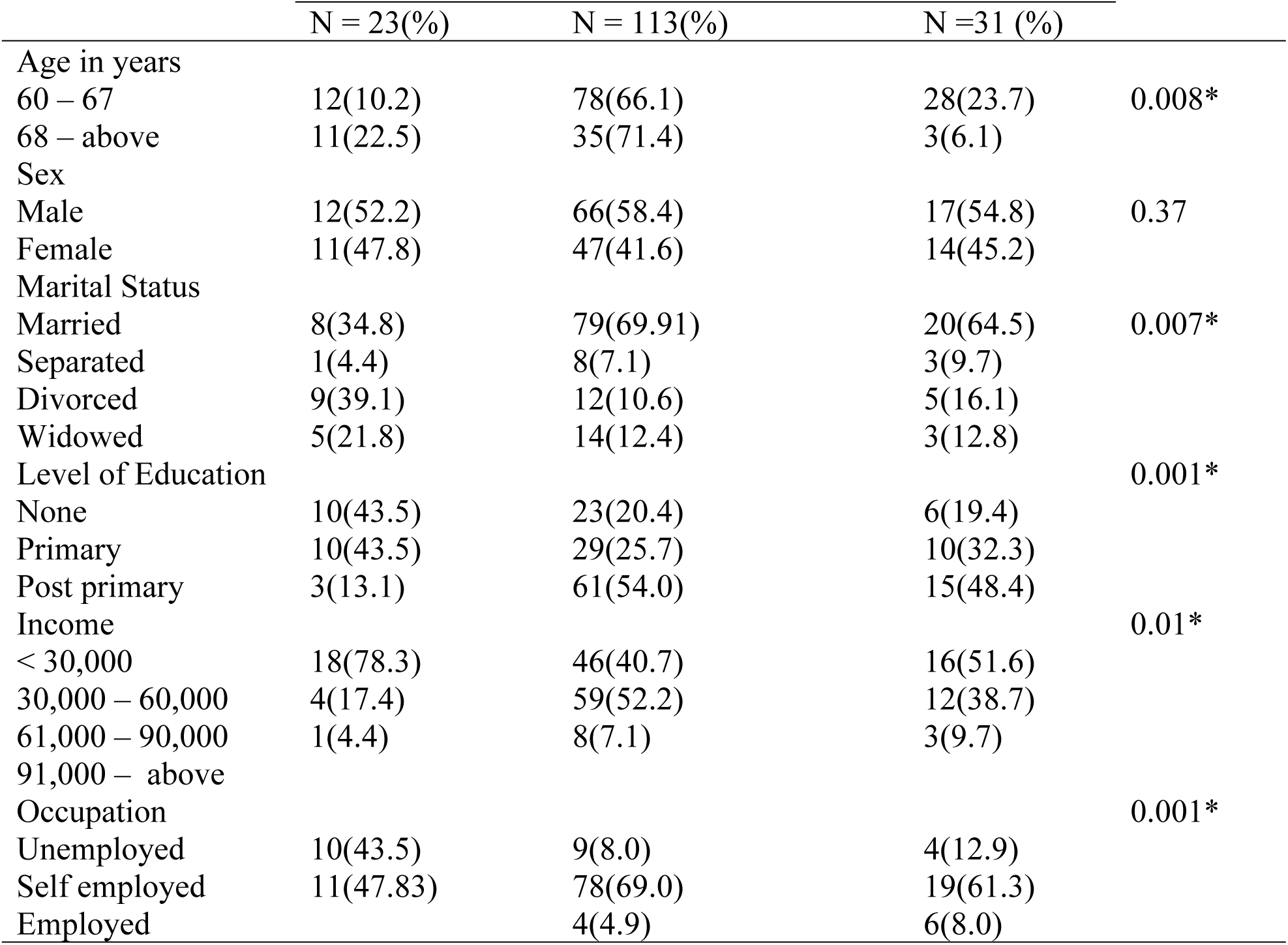
Association between socio - demographic variables and nutritional status of respondents in rural area.

Table 6 revealed that there is a statistically significant association between the nutritional status and some socio demographic (Age, ethnicity and occupation) of the respondents in urban area of the participants with P-Value of 0.001,0.01 and 0.007 respectively. Those who aged between 68 years and above were more at risk of malnutrition this related in compare with rural area but those who are unemployed were more malnourished which what was obtained in rural area is contrary to this.

**Table 6:** Association between socio - demographic variables and nutritional status of participants in urban area.

| Variable | Nutritional Status |  |  | P – value |
| --- | --- | --- | --- | --- |
|  | Malnourished<br>N = 8(%) | At risk of malnutrition<br>N = 81(%) | Normal<br>N =75 (%) |  |
| Age in years |  |  |  |  |
| 60 – 67 | 2(2.0) | 43(42.2) | 57(55.9) | 0.001* |
| 68 – above | 6(9.7) | 38(61.3) | 18(29.0) |  |
| Ethnicity |  |  |  | 0.01* |
| Igbo | - | 6(7.4) | 11(14.7) |  |
| Yoruba | 7(87.5) | 74(91.4) | 64(85.3) |  |
| Other | 1(12.5) | 1(1.2) | - |  |
| Occupation |  |  |  | 0.007* |
| Unemployed | 7(87.5) | 29(35.8) | 18(24.0) |  |
| Self employed | 1(12.5) | 48(59.3) | 51(68.0) |  |
| Employed |  | 4(4.9) | 6(8.0) |  |

\* Statistically significant p – value < 0.05
Pearsons's chi - square

In table 7, All the associated factors which showed a statistically significant association with nutritional status in the univariable regression analysis at p-value = 0.2 and the one considered APIORI were put in a multiple logistic regression model to assess individual variable effect removing any possible effects of confounders using a backward elimination method. Following the regression analysis, age, quality of life and family support were found to be significant predictor of nutritional status among the various factors entered into the model. Age (P < 0.002), quality of life (P < 0.003) and family support (P < 0.01). Those with strong family support were less likely to be malnourished with odd ratio of 0.3 ( 95%CI:0.11 – 0.82) than those who have No family support and those with good quality of life were less likely to be malnourished or risk of malnutrition with an odd ratio of 0.2 (95% CI: 0.07 – 0.58) than those with poor quality of life

**Table 7:** Logistic regression of the predictors of nutritional status in rural area.

| Variable | Univariable |  |  |  | Multivariable |  |  |  |
| --- | --- | --- | --- | --- | --- | --- | --- | --- |
|  | OR | 95% CI |  | P-value | AdjOR | 95% CI |  | P-value |
|  |  | Lower | Upper |  |  | Lower | Upper |  |
| Age | 1.1 | 1.1 | 1.2 | 0.001* | 1.1 | 1.0 | 1.2 | 0.002* |
| Family support |  |  |  |  |  |  |  |  |
| No | 1.0 | Ref | Ref | Ref | 1.0 | Ref | Ref | Ref |
| Weak | -1.62 | -3.69 | 0.44 | 0.13 | 0.1 | 0.01 | 1.31 | 0.07 |
| Strong | -1.22 | -3.07 | -0.44 | 0.002 | 0.3 | 0.11 | 0.82 | 0.01* |
| Quality of life |  |  |  |  |  |  |  |  |
| Poor | 1.0 | Ref | ref | Ref | 1.0 | Ref | Ref | Ref |
| Good | 0.13 | 0.05 | 0.32 | 0.001* | 0.2 | 0.07 | 0.58 | 0.003* |
| Sex |  |  |  |  |  |  |  |  |
| Female | 1.0 | Ref |  | Ref | 1.0 | Ref | Ref | Ref |
| Male | 0.96 | 0.51 | 1.84 | 0.91 | 0.7 | 0.3 | 1.4 | 0.27 |
| Marital status |  |  |  |  |  |  |  |  |
| Married | 0.33 | 0.12 | 0.87 | 0.03 | 0.84 | 0.29 | 2.43 | 0.75 |
| Divorced | 1.0 | Ref | Ref | Ref | 1.0 | Ref | Ref | Ref |
| Separated | 0.26 | 0.06 | 1.17 | 0.08 | 0.50 | 0.09 | 2.68 | 0.43 |
| Widowed | 0.88 | 0.26 | 3.07 | 0.85 | 1.13 | 0.28 | 4.51 | 0.86 |
| Education |  |  |  |  |  |  |  |  |
| None | 1.0 | Ref | Ref | Ref | 1.0 | Ref | Ref | Ref |
| Primary | 0.65 | 0.26 | 1.66 | 0.37 | 0.98 | 0.37 | 2.67 | 0.98 |
| Post primary | 0.42 | 0.18 | 0.98 | 0.05 | 0.87 | 0.31 | 2.49 | 0.81 |
| Income |  |  |  |  |  |  |  |  |
| < 30,000 | 1.0 | Ref | Ref | Ref | 1.0 | Ref | Ref | Ref |
| 31,000 – 60, 000 | 0.62 | 0.32 | 1.23 | 0.17 | 1.4 | 0.62 | 3.32 | 0.34 |
| 61,000 – 90,000 | 0.49 | 0.14 | 1.73 | 0.27 | 1.5 | 0.38 | 6.50 | 0.51 |

\* Statistically significant $p < 0.05$ AdjOR. = Adjusted; OR = Odds ratio; CI = Confidence interval

In table 8, there is an increase in number of variables found to be significant predictor of nutritional status among the various factors entered into the model in compare with rural area. Age (OR: 1.1, 95%CI: 1.01 – 1.16, P < 0.01), quality of life (OR: 0.29, 95%CI: 0.1 – 0.9, P < 0.03), sex (OR: 3.3, 95%CI: 1.04 – 10.5, P < 0.01), education level (OR: 2.05, 95%CI: 1.02 – 4.16, P < 0.04), and income (OR:0.21, 95%CI: 0.04 – 0.93, P < 0.04) were found to be significant. Those with good family support were less likely to be malnourished with odd ratio of 0.29 ( 95%CI:0.1 – 0.9) than those who have low quality of life. This correlated with what was obtained in rural area but family support is not among the predictors nutritional status in urban area.

**Table 8:** Logistic regression of the predictors of nutritional status in urban area.

| Variable | Univariable |  |  |  | Multivariable |  |  |  |
| --- | --- | --- | --- | --- | --- | --- | --- | --- |
|  | OR | 95% CL |  | P-value | AdjOR | 95% CL |  | P-value |
|  |  | Lower | Upper |  |  | Lower | Upper |  |
| Age | 1.10 | 1.03 | 1.17 | 0.002* | 1.1 | 1.01 | 1.16 | 0.01* |
| Family support |  |  |  |  |  |  |  |  |
| No | 1.0 | Ref | Ref | Ref | 1.0 | Ref | Ref | Ref |
| Weak | -1.22 | -2.47 | 0.31 | 0.06 | 0.38 | 0.09 | 1.48 | 0.16 |
| Strong | -1.42 | -2.49 | -0.34 | 0.01* | 0.39 | 0.11 | 1.37 | 0.15 |
| Quality of life |  |  |  |  |  |  |  |  |
| Poor | 1.0 | Ref | Ref | Ref | 1.0 | Ref | Ref | Ref |
| Good | 0.19 | 0.07 | 0.47 | 0.001* | 0.29 | 0.1 | 0.9 | 0.027* |
| Sex |  |  |  |  |  |  |  |  |
| Female | 1.00 | Ref | Ref | Ref | 1.0 | Ref | Ref | Ref |
| Male | 0.99 | 0.48 | 1.99 | 0.97 | 3.3 | 1.04 | 10.5 | 0.04* |
| Marital status |  |  |  |  |  |  |  |  |
| Married | 0.85 | 0.32 | 2.29 | 0.76 | 0.7 | 0.23 | 2.32 | 0.61 |
| Divorced | 1.0 | Ref | Ref | Ref | 1.0 | Ref | Ref | Ref |
| Separated | 2.19 | 0.57 | 8.43 | 0.25 | 1.7 | 0.41 | 7.54 | 0.44 |
| Widowed | 1.28 | 0.45 | 3.68 | 0.64 | 0.99 | 0.31 | 3.17 | 0.99 |
| Education |  |  |  |  |  |  |  |  |
| None | 1.0 | Ref | Ref | Ref | 1.0 | Ref | Ref | Ref |
| Primary | 1.47 | 0.78 | 2.76 | 0.23 | 2.05 | 1.02 | 4.16 | 0.04* |
| Post primary | 0.79 | 0.23 | 2.79 | 0.73 | 1.43 | 0.36 | 5.65 | 0.61 |
| Income |  |  |  |  |  |  |  |  |
| < 30,000 | 1.0 | Ref | Ref | Ref | 1.0 | Ref | Ref | Ref |
| 31,000 – 60, 000 | 0.61 | 0.32 | 1.16 | 0.04* | 0.54 | 0.25 | 1.16 | 0.11 |
| 61,000 – 90,000 | 0.33 | 0.11 | 0.93 | 0.13 | 0.21 | 0.04 | 0.93 | 0.04* |

\* Statistically significant p<0.05 AdjOR. = Adjusted; OR = Odds ratio; CL = Confidence interval

According to table 9,There is a statistically significant association between the quality of life score of the participants and their place of residence (P = 0.01)

**Table 9:**
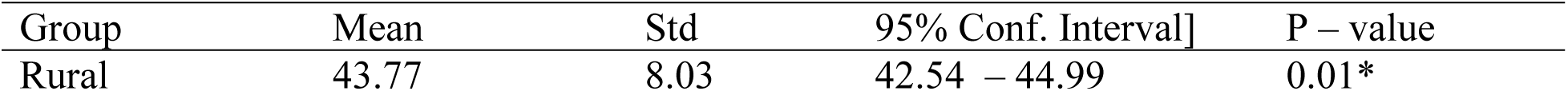
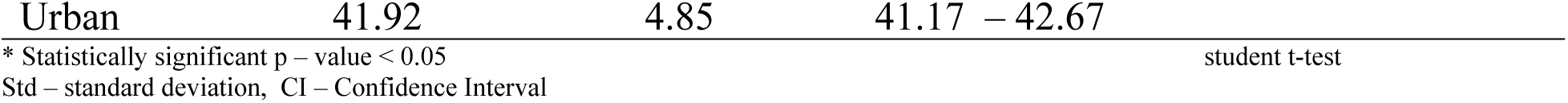
Association between quality of life score and place of residence.

| Group | Mean | Std | 95% Conf. Interval] | P – value |
| --- | --- | --- | --- | --- |
| Rural | 43.77 | 8.03 | 42.54 – 44.99 | 0.01* |

|  |  |  |  |
| --- | --- | --- | --- |
| Urban | 41.92 | 4.85 | 41.17 – 42.67 |
| * Statistically significant p – value < 0.05 |  |  | student t-test |
| Std – standard deviation, CI – Confidence Interval |  |  |  |

According table 10 below, there is a significant association between nutritional status and marital status, level of education, income and occupation of participants in rural area with p-value of 0.001, 0.001, 0.003, and 0.001 respectively. Poor quality of life is higher among male participants and poor quality of life is higher among the unemployed. There is a significant association between nutritional status and marital status, income and occupation of participants in urban area with p-value of 0.001, 0.001 and 0.001respectively. This is similar to what obtained in rural area, while age, sex and ethnicity were observed to have significant association with nutritional status in Urban area with p-value of 0.02, 0.03 and 0.015 respectively. Participants aged 68 – above have the higher percentage of poor quality of life and male gender also have higher percentage of poor quality of life.

**Table 10:** Association between socio - demographic variables and quality of life (QoL) of the participants in rural and urban area.

| Variables | Quality of Life |  |  |  |  |  |
| --- | --- | --- | --- | --- | --- | --- |
|  | Rural |  |  | Urban |  |  |
|  | Poor QoL | Good QoL | P-value | Poor QoL | Good QoL | P-value |
| Age in years |  |  |  |  |  |  |
| 60 – 67 | 28(23.8) | 90(76.27) | 0.35 | 14(13.73) | 88(86.27) | 0.02* |
| 68 – above | 15(30.6) | 34(69.39) |  | 18(29.03) | 44(70.97) |  |
| Sex |  |  |  |  |  |  |
| Male | 28(29.5) | 67(70.53) | 0.2 | 3(7.89) | 35(92.11) | 0.03* |
| Female | 15(20.8) | 57(79.17) |  | 29(23.02) | 97(76.98) |  |
| Marital Status |  |  | 0.001* |  |  | 0.009* |
| Married | 18(16.8) | 89(83.18) |  | 9(11.11) | 72(88.89) |  |
| Separated | 6(50.0) | 6(50) |  | 4(28.57) | 10(71.43) |  |
| Divorced | 14(53.9) | 12(46.15) |  | 2(10) | 18(90.00) |  |
| Widowed | 5(23.8) | 17(77.19) |  | 17(35.42) | 32(65.58) |  |
| Ethnicity |  |  | 0.3 |  |  | 0.015* |
| Igbo | 5(31.3) | 11(68.75) |  | 3(17.65) | 14(82.35) |  |
| Yoruba | 18(20.7) | 69(79.31) |  | 27(18.62) | 118(81.38) |  |
| Other | 20(29.8) | 44(70.18) |  | 2(100) |  |  |
| Level of Education |  |  | 0.001* |  |  | 0.06 |
| None | 20(51.28) | 19(48.72) |  | 18(26.87) | 49(73.13) |  |
| Primary | 14(28.57) | 35(71.43) |  | 14(16.28) | 72(83.72) |  |
| Post primary | 9(10.96) | 70(89.04) |  |  | 11(100) |  |
| Income |  |  | 0.003* |  |  | 0.001* |
| < 30,000 | 30(37.50) | 50(62.50) |  | 24(32.43) | 50(67.57) |  |
| 30,000 – 60,000 | 12(16) | 63(84) |  | 8(11.27) | 63(88.73) |  |
| 61,000 – 90,000 | 1(8.33) | 11(91.67) |  |  | 19(100) |  |
| 91,000 – above |  |  |  |  |  |  |
| Occupation |  |  | 0.001* |  |  | 0.001* |
| Unemployed | 14(60.87) | 9(39.13) |  | 20(37.04) | 34(62.96) |  |
| Self employed | 22(20.37) | 86(79.63) |  | 11(11.00) | 89(89.00) |  |
| Employed | 7(19.44) | 29(80.56) |  | 1(10) | 9(90) |  |

In table 11, all the associated factors which showed a statistically significant association with nutritional status in the univariable regression analysis at p-value = 0.2 and the one considered APIORI were put in a multiple logistic regression model to assess individual variable effect removing any possible effects of confounders using backward elimination method. Following the regression analysis, age, quality of life and family support were found to be significant predictor of nutritional status among the various factors entered into the model. Family support (P = 0.002, 95%CI=2.03 – 20.50), sex (P = 0.02, 95%CI=0.04 – 0.72), marital status (P = 0.02, 95%CI=1.20 – 20.4) and nutritional status (P= 0.002, 95%CI=3.24 – 175.74). Those with strong family support were more likely to have good quality of life with odd ratio of 2.03 (P = 0.002, 95%CI=2.03 – 20.50) than those who have No family support and those with normal nutritional status were more likely to have good quality of life in 28 times than those that were malnourished (P= 0.002, 95%CI=3.24 – 175.74).

**Table 11:** Predictors of quality of life in rural area.

| Variable | Quality of life |  |  |  |  |  |  |  |
| --- | --- | --- | --- | --- | --- | --- | --- | --- |
|  | Univariable |  |  |  | Multivariable |  |  |  |
|  | OR | 95% CI |  | P-value | AdjOR | 95% CI |  | P-value |
|  |  | Lower | Upper |  |  | Lower | Upper |  |
| Family support |  |  |  |  |  |  |  |  |
| No | 1.0 | Ref | Ref | Ref | 1.0 | Ref | Ref | Ref |
| Weak | 0.76 | 0.11 | 5.01 | 0.77 | 0.1 | 0.08 | 16.58 | 0.8 |
| Strong | 7.21 | 3.28 | 15.86 | 0.001* | 0.3 | 2.03 | 20.50 | 0.002* |
| Sex |  |  |  |  |  |  |  |  |
| Female | 1.0 | Ref |  | Ref | 1.0 | Ref | Ref | Ref |
| Male | 0.96 | 0.51 | 1.84 | 0.91 | 0.2 | 0.04 | 0.72 | 0.02* |
| Marital status |  |  |  |  |  |  |  |  |
| Married | 5.77 | 2.29 | 14.51 | 0.001* | 4.96 | 1.20 | 20.4 | 0.02* |
| Divorced | 1.0 | Ref | Ref | Ref | 1.0 | Ref | Ref | Ref |
| Separated | 1.16 | 0.29 | 4.58 | 0.8 | 0.2 | 0.03 | 2.11 | 0.2 |
| Widowed | 3.73 | 1.05 | 13.24 | 0.04* | 3.67 | 0.59 | 22.71 | 0.16 |
| Income |  |  |  |  |  |  |  |  |
| < 30,000 | 1.0 | Ref | Ref | Ref | 1.0 | Ref | Ref | Ref |
| 31,000 – 60, 000 | 3.15 | 1.46 | 6.77 | 0.003* | 1.2 | 0.38 | 3.59 | 0.77 |
| 61,000 – 90,000 | 6.6 | 0.81 | 53.71 | 0.08 | 1.5 | 0.23 | 24.98 | 0.77 |
| Nutrition status |  |  |  |  |  |  |  |  |
| Malnourished | 1.0 | Ref | Ref | Ref | 1.0 | Ref | Ref | Ref |
| Risk of malnutrition | 43.55 | 11.52 | 164.6 | 0.001* | 74.99 | 11.61 | 484.45 | 0.001* |
| Normal | 19.16 | 4.46 | 82.19 | 0.001* | 23.84 | 3.24 | 175.74 | 0.002* |

In table 12, all the associated factors which showed a statistically significant association with nutritional status in the univariable regression analysis at p-value = 0.2 and the one considered APIORI were put in a multiple logistic regression model to assess individual variable effect removing any possible effects of confounders using backward elimination method. Following the regression analysis, family support, marital status and income were found to be significant predictor of quality of life among the various factors entered into the model. Family support (P = 0.001, 95%CI=6.59 – 234.29), marital status (P = 0.01, 95%CI=0.002 –0.51) and income (P= 0.02, 95%CI= 1.32 – 16.82). Those with strong family support were more likely to have good quality of life with odd ratio of 15.5 (P = 0.001, 95%CI=6.59 – 234.29) than those who have No family support

**Table 12:**
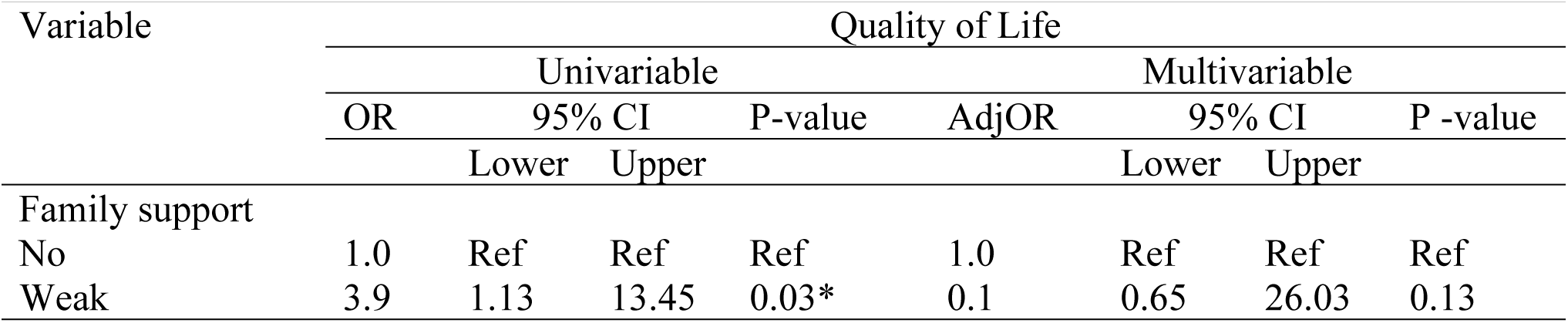
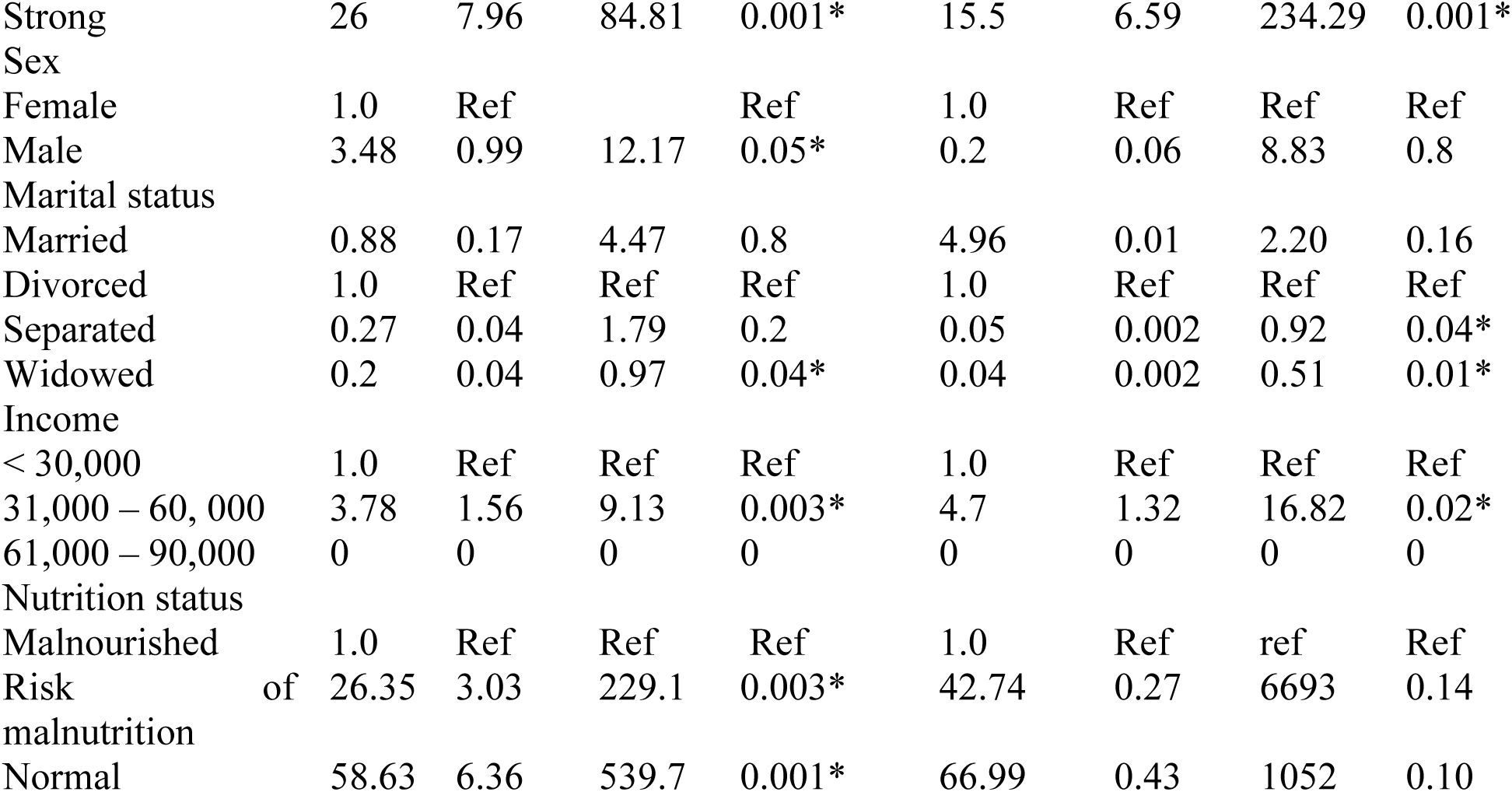
Predictor of quality of life in urban area.

In table 13, there is a significant relationship between the nutritional status and quality of life in both rural and urban area with p – value of 0.001 and 0.001 respectively. Majority those with poor quality of life in rural area were malnourished while a little bit above average of those who have poor quality of life in urban area were at risk of malnutrition

**Table 13:**
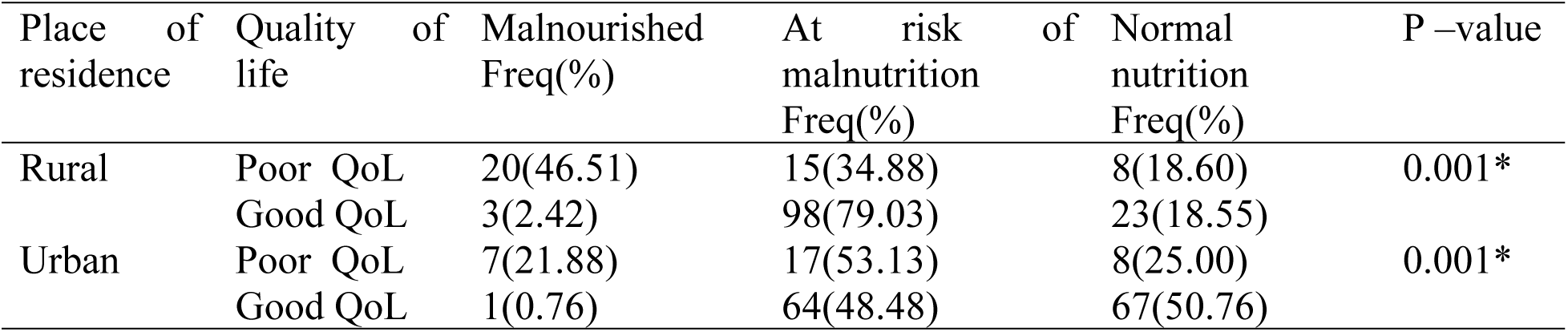
Association between nutritional status and quality of life of participants in Rural and Urban area.

| Place of residence | Quality of life | Malnourished Freq(%) | At risk of malnutrition Freq(%) | Normal nutrition Freq(%) | P –value |
| --- | --- | --- | --- | --- | --- |
| Rural | Poor QoL | 20(46.51) | 15(34.88) | 8(18.60) | 0.001* |
|  | Good QoL | 3(2.42) | 98(79.03) | 23(18.55) |  |
| Urban | Poor QoL | 7(21.88) | 17(53.13) | 8(25.00) | 0.001* |
|  | Good QoL | 1(0.76) | 64(48.48) | 67(50.76) |  |

## DISCUSSION

The majority of the participants in rural and urban area falls within the age of 60 – 67 years and the mean age were 65.4±5.5 and 66.1±5.1 respectively for rural and urban area. This is similar to what was reported by Sabita et al.[28]This was a little bit lower to what was reported in a study among elderly in Niger.[10] This may be as a result of higher life expectancy in Niger than Nigeria.[29] More than half of the participants (56.87%) in rural area were male. This was closed to what was reported in study among elderly in Nepal [28] and India [30] while most of the participants (76.83%) in urban area where female. This correlate with what was reported by different studies.[4,31,32] Larger percentage of the participants were married in both rural and urban area. The participants are predominantly Yoruba in both rural and urban area but it was observed that significant number of participants (34.13%) in rural area were Egun this is as a result of a larger population of Egun people in Badagry. It was revealed that Christianity is the religion most practiced by the participants in both rural and urban area. The percentage of participants with less than 30,000 monthly income in both rural (47.90%) and urban(45.12%) was a little bit close to the percentage of participants with 61,000 – 90,000 monthly income in both rural (44.91%) and urban (43.29)%. This is similar to what was reported by Alao et al.^10^ Majority participants in rural and urban area were self employed. The mean weight of the participants in rural and urban area was 63.91±7.78kg and 60.21±12.43 respectively this was close to what was reported by Afolabi et al in a study among elderly in southwest Nigeria.[6] Using BMI, 35.45% of the participants in rural area were either underweight or overweight while 29.89% of the participants in urban area were either underweight or overweight. Level of malnutrition in rural area is higher than urban area as reported by different studies.[11,33] The mean BMI was found to be 23.73±3.08kg/m^2^ in rural and 23.04±4.27k g/m^2^ in urban. This is a little bit close to what was reported by Aganiba et al.[33]Female who are underweight have less mean score of BMI in rural while in urban area female who are underweight have high mean score of BMI. Assessment of the nutritional status using Mini nutritional assessment short form (MNA - SF) revealed that among rural dwellers 13.77% were malnourished, 67.66% were at risk of malnutrition and 18.6% were well nourished. This is similar to what was reported by Dewi et al in a study done in rural area of India [9] Although this was a little bit different with what was reported by Kirtana in which 57.4% were at risk of malnutrition and this different may be as a result of better condition of living of elderly who participated in the study. In urban area, the participants that are well nourished are higher than what obtained in the rural area. 45.73% were well nourished, 49.39% were at risk of malnutrition and 4.88% were malnourished. This findings was similar to the findings of several studies among urban dwellers elderly. [11,34]. The risk of malnutrition among the elderly in the rural and urban area is so alarming, majority of the participants in both rural and urban area where at risk of malnutrition using MNA – SF assessment but risk of malnutrition was among rural dwellers is higher than urban dwellers. 81.43% of participants in rural area is either malnourished or at risk of malnutrition which is higher than what was reported in various studies [9, 11,34]. Also, 54.27% of the participants were either malnourished or at risk of malnutrition this was similar to the findings of several studies. But it was observed that using BMI majority of the respondents have normal nutrition. There are reports that undernutrition and overnutrition encompasses dietary history, physical (including anthropometry), clinical, psychological and self assessment of nutritional state and not anthropometric measurements of BMI alone. Also, there had been many controversies on the measurement of the true height of the elderly due to the reduction invertebral height with ageing. This led some author to propose knee length as a proxy for the height of older people.[30]

A statistically significant association was observed between nutritional status and age group, marital status, level of education, income and occupation among the rural dwellers with P-Value of 0.008, 0.007, 0.001, 0.01 and 0.001 respectively. This is similar to what was reported in a study among geriatric population in Nepal.[18,30] Adebusoye et al also reported a significant association between nutritional status and some socio demographic (age, marital status and occupation).[8] Baweja and Wadhwa in their different studies reported that only age has significant association with nutritional status and was reported that been risk of malnutrition increase by age.[35,36]but there is no significant association between nutritional status and sex, ethnicity and religion this is similar to what was found by Ghimire et al[37]. This may be as a result of more number of people from same group and religion. People aged 68 and above have higher level of malnutrition this was also reported by a study. Also, those that were divorced have higher percentage of malnutrition and those with less income. This may be as a result of not financially stable.

In urban dwellers, a statistically significant association was observed between nutritional status and age, ethnicity and income and others socio demography did not have association with the nutritional status. This was similar to what was reported in a study conducted in Niamey.[10]

The study conducted in Pharping, Nepal[30] Spain[38], and Iran[39] showed that people with poor economic status and educational status were at higher risk of being malnourished. This is the fact which shows that socioeconomic and educational difference affects the nutritional status of individuals. Similarly, past occupations of the elders also affect their nutritional status. In rural dwellers, Age (P < 0.002), quality of life (P < 0.003) and family support (P < 0.01) were observed to be the predictor of malnutrition among elderly. The level of been malnourished increases by more than one fold of age. (95%CI: 1.0 – 1.2).Those with strong family support were less likely to be malnourished with odd ratio of 0.3 ( 95%CI:0.11 – 0.82) than those who have No family support and those with good quality of life were less likely to be malnourished or risk of malnutrition with odd ratio of 0.2 (95% CI: 0.07 – 0.58) than those with poor quality of life. This is similar to what was reported in a study conducted in Ethiopia.[11] Also, Sabita et al only reported age as the predictor of nutritional status in the study among elderly in Nepal. In urban dwellers, there is an increase in the number of predictors of nutritional status when compared with rural area. Age (OR: 1.1, 95%CI: 1.01 – 1.16, P < 0.01), quality of life (OR: 0.29, 95%CI: 0.1 – 0.9, P < 0.03), sex (OR: 3.3, 95%CI: 1.04 – 10.5, P < 0.01), education level (OR: 2.05, 95%CI: 1.02 – 4.16, P < 0.04),and income (OR:0.21, 95%CI: 0.04 – 0.93, P < 0.04) were found to be significant. This revelation is similar to what was reported by different studies. [8,11,30] Male are likely to be more malnourished than female in three fold (95%CI: 1.04 – 10.5). Majority of participants in both rural and urban have god quality of life. 25.75% of the participants in rural area have poor quality of life while 19.51% have poor quality of life in urban area. This is far better to what was reported by several studies. Dewi et al reported that more than half of elderly in rural area have low QoL.[9] Also, Ning et al reported low QoL among rural dwellers elderly in China in which age, economic status, living conditions and level of education showed statistically significant association with QoL.[40] This may be as a result of the living conditions of elderly participated in the study. But Oladipupo et al reported a similar value in the study among elderly in southwestern of Nigeria.[4]

In comparing the mean score of quality of life with the place of residence a significant association was observed with p – value 0.01. The mean score was higher in rural area (43.77±8.03) than urban area (41.93±4.85). This is a little bit higher than what was reported by Sabita et al[11] and a little bit less than what was reported in a study in Iran.[41]The differences in sample population and sample sizes and data collection tools may be attributed as the reasons for the differing figures.

There is a significant association between nutritional status and some socio demography (marital status, income and occupation) of participants in both rural and urban area while level of education is only significant in rural area and age, sex and ethnicity were observed to have significant association with nutritional status in urban area. Ning et al reported something similar in the study among rural dwellers elderly in China in which age, economic status, living conditions and level of education showed statistically significant association with QoL. Also, A study of QoL among elderly in rural populace in Nigeria reported that over average have good quality of life and age, gender, marital status, occupation and income were statistically significant with the QoL. [12] Sabita et al also reported that different socio demographic factors such as age, ethnicity, religion, marital status, educational status, and past occupation are positively associated with an individual’s quality of life.[30] Gureje et al reported that only age has significant relationship with quality of life. This difference may be as a result of different in sample size. In rural, Family support (P = 0.002, 95%CI=2.03 – 20.50), sex (P = 0.02, 95%CI=0.04 – 0.72), marital status (P = 0.02, 95%CI=1.20 – 20.4) and nutritional status (P= 0.002, 95%CI=3.24 – 175.74). Those with strong family support were more likely to have good quality of life with odd ratio of 2.03 (P = 0.002, 95%CI=2.03 – 20.50) than those who have No family support and those with normal nutritional status were more likely to have good quality of life in 28 times than those that were malnourished (P= 0.002, 95%CI=3.24 – 175.74). This is similar to what was reported in a study in Nigeria among rural populace elderly[42] but only gender was reported to be predictor of quality of life in a study Iran.[20].In urban dwellers, family support, marital status and income were found to be significant predictor of quality of life among the various factors entered into the model. Family support (P =, 95%CI=6.59 – 234.29), marital status (P = 0.01, 95%CI=0.002 –0.51) and income (P= 0.02, 95%CI= 1.32 – 16.82). Those with strong family support were more likely to have good quality of life with odd ratio of 15.5 (P = 0.001, 95%CI=6.59 – 234.29) than those who have No family support. This was a little bit different with what was reported by Sabita et al in the study among urban dweller in Nepal.[30] This may due to the different in population size. A significant association was found between nutritional status and quality of life in both rural and urban area with p - value 0.001 and 0.001 respectively. This underline the importance of considering malnutrition when attempting to improve QOL. Dewi et al reported a significant association at p<0.05 between nutritional status and quality of life among elderly in rural area which further revealed that those with poor quality of life have higher risk of malnutrition.[9] A significant association between QoL and nutritional status was also reported by Eleni.[42] Sabita et al also reported a significant association between nutritional status and quality of life among geriatric population in Nepal.[30]

## Conclusion and recommendation

The study assessed the nutritional status and quality of life of the elderly in the rural and urban area. The majority of the participants in the rural area were at risk of malnutrition while close to half of the participants in rural area were at risk of malnutrition. The percentage of malnourished is high among male gender in rural area while is high among female gender in urban area.

Predictors of malnutrition includes age, family support, quality of life, income, level of education and occupation status while family support, nutritional status, marital status, income and sex were found to be predictors of quality of life. To tackle malnutrition, Individual: Elderly should ensure that they eat adequate diet and make it as a routine to always check their nutritional status

### Strength and limitation of study

Strength of the study: This study was analysed using STATA which is one of the best tool for data analysis in health research. Also, it is a large comparative study in rural and urban area and strategies for nutritional and quality of life implementation are unique for primary care.

#### Limitation of the study

This study had some limitations. It is a cross sectional study not causal relationship. Also, nutritional status is subjective from recall and when necessary the care giver is asked to provide the information. It cannot be generalized because the study was done in at a single site using a moderate sample size

#### Funding

No external source of funding

#### Authors Contributions

Conceptualization: Babatunde Abdulmajeed Akodu, Monsuru Owolabi Badmus

Data curation: Monsuru Owolabi Badmus, Babatunde Abdulmajeed Akodu, Moninuola Seliat Ojikutu, Taiwo Hussain Agunbiade

Formal analysis: Monsuru Owolabi Badmus, Gbenga Olorunfemi, Babatunde Abdulmajeed Akodu, Olufunmilayo Abeni Olokodana-Adesalu

Methodology: Babatunde Abdulmajeed Akodu, Monsuru Owolabi Badmus, Gbenga Olorunfemi, Oluchi Joan Kanma-Okafor,

Project administration: Babatiunde Abdulmajeed Akodu, Monsuru Owolabi Badmus, Oluchi Joan Kanma-Okafor

Supervision: Babatunde Abdulmajeed Akodu, Oluchi Joan Kanma-Okafor, Gbenga Olorunfemi, Olufunmilayo Abeni Olokodana-Adesalu

Validation: Babatunde Abdulmajeed Akodu, Gbenga Olorunfemi, Oluchi Joan Kanma-Okafor,

Visualization: Babatunde Abdulmajeed Akodu, Gbenga Olorunfemi, Seliat Ojikutu, Taiwo Hussain Agunbiade

Writing - original draft: Babatunde Abdulmajeed Akodu

Writing – review and editing: Babatunde Abdulmajeed Akodu, Gbenga Olorunfemi, Oluchi Joan Kanma-Okafor, Olufunmilayo Abeni Olokodana-Adesalu

## Data Availability

All relevant data are within the manuscript and its Supporting Information files.

